# Fathers’ and Mothers’ Support Needs and Support Experiences after Rapid Genome Sequencing

**DOI:** 10.1101/2025.07.01.25330667

**Authors:** Helen Dolling, Sophie Rowitch, Malachy Bromham, Stephanie Archer, Sara O’Curry, David H. Rowitch, F. Lucy Raymond, Claire Hughes, Kate Baker

**Author notes:** **Corresponding author:** Helen Dolling, Centre for Child, Adolescent, and Family Research, Department of Psychology, University of Cambridge, Free School Lane, Cambridge, CB2 3RQ, UK, E.

## Abstract

As early rapid genomic sequencing (rGS) is adopted in paediatric medicine, there is an urgency to understand and address family support needs. This mixed methods study (*Peregrin\**) examined the experiences of 96 parents, 1–5 years after receiving trio rGS results for their child with a severe early-onset condition. Quantitative outcome measures assessed parental well-being, life satisfaction, and family impact, comparing results to non-clinical population data, between mothers and fathers, and according to child’s diagnostic outcome. Qualitative semi-structured interviews explored parents’ satisfaction with support, engagement with support networks, and unmet needs. Quantitatively, mothers exhibited elevated anxiety and depression relative to population norms, and there was a lack of strong correlation in well-being metrics within couples. Parents of children with a genomic diagnosis reported poorer well-being, explained by greater medical complexity. Qualitatively, insufficient support was more frequently reported by those whose child had received a genomic diagnosis (36%) compared to those without (6%). Families drew on a range of formal and informal support sources, including condition-specific groups, though these were accessed by a minority of fathers. These findings highlight persistent and evolving support needs in families affected by complex childhood health conditions, which persist after rGS. Parents’ support needs are highly individual, vary over time and across children’s illness trajectory. There remain important gaps between parental needs and support, impacting on family well-being.

## INTRODUCTION

Genomic testing is becoming available in healthcare settings worldwide; rapid genome sequencing (rGS) is now offered in neonatal and paediatric intensive care units (NICU/PICU) in England. More families are receiving diagnoses of rare or ultra-rare conditions, many being life-threatening or life-limiting, early in their child’s life (1). Early genomic testing and diagnosis can have clinical and personal benefits (2,3), but can also cause anxiety, distress, uncertainty and upheaval (4,5). Studies of parental support needs in the context of paediatric rare diseases and genomic diagnostics remain scarce (6,7).

Having a child admitted to a NICU/PICU with an unexpected severe illness can be traumatic, with long-term implications for families (8,9). A range of risk and protective factors explain why some families adjust and adapt, while others struggle to maintain family functioning. Factors include problem-solving and coping styles, family relationships, socioeconomic status (SES) and resource availability (10,11). Caregiver well-being is also influenced by a child’s medical complexity, adaptive functioning and behaviour (12–14). Support is consistently identified as a protective factor for family adjustment and, as a modifiable factor, provides a natural focus for interventions (15).

As genomic testing becomes more common, it is important to understand the perspectives of caregivers regarding the need for and availability of support. The transition to parenthood, irrespective of a child’s health, is often accompanied by reductions in parental relationship quality and mutual support (18,19). Parents differ in their responses to childhood illness, with mothers reporting higher levels of anxiety, depression and stress symptoms than fathers (20,21). Previous research has highlighted possible gender and cultural differences in values and norms around stress, coping, and the role of support (22,23). Disease rarity may constrain support availability and amplify psychosocial impacts of childhood-onset rare disease on parents (24).

To address these issues, the Peregrin* (Parental Experiences of Rapid Early Genomic Results in Infancy) study followed-up parents whose children had received rGS because of early-onset illness. The first objective was to evaluate parents’ psychosocial needs, using validated questionnaires. We compared parental well-being, life satisfaction and family impact with non-clinical population data, between mothers and fathers, and according to presence / absence of a genomic diagnosis. The second objective was to explore parents’ support experiences through qualitative interviews, focusing on perceptions and sources of support, and attitudes to support groups.

Support is a multidimensional construct that includes emotional, informational and instrumental elements, and encompasses both actual and perceived support (16). Adopting Cohen’s definition of support as “*a social network’s provision of psychological and material resources intended to benefit an individual’s ability to cope with stress*” (17), we applied a multi-layered approach incorporating functional (i.e. perceived availability of support) and structural (i.e. extent, diversity and frequency of interactions) components. By integrating these objectives and approaches, unmet support needs can be identified, and recommendations drawn.

## MATERIALS AND METHODS

### Participants

This article reports a subset of data from Peregrin*, a follow-up study of 96 parents of infants and children who had received rGS within the Next Generation Children project (NGC) (25,26). NGC was a translational research study (November 2016 – August 2020) with broad inclusion criteria to explore feasibility, acceptability, clinical utility, and logistics of implementing rGS in NICU/PICU within tertiary level hospitals in the UK. Families (n = 521, 90% trios) were recruited from three NICU, two PICU, and by rapid referrals from Paediatric Neurology and Clinical Genetics in the East and Southeast of England. The cohort reflected the distribution of SES within the populations served by the recruiting hospitals. Consent was provided for long-term follow-up via NIHR Rare Diseases BioResource. The Peregrin* cohort (data collected December 2021 – January 2023) thus represents the first large group of UK-based infants and children with any severe, complex, and unexplained symptoms to receive early rGS in hospital settings, before NHS implementation.

Parents were eligible for Peregrin* if they had participated in NGC and they were a parent of (a) an infant (≤2 years at rGS), whether or not a genomic diagnosis had been received; or (b) a child (>2 years at rGS), who had received a genomic diagnosis. Parents whose child met the above criteria and had died > 12 months previously were eligible. Parents of children >2 years at rGS without a genomic diagnosis were excluded because their experiences are represented within existing literature on diagnostic odyssey (27,28).

Recruitment followed a cluster stratification approach (29), with child’s age (≤2 years or >2 years at rGS) as the stratification variable and Genomic Diagnosis/No Genomic Diagnosis as cluster boundaries. We aimed to recruit 30 families per cluster to enable subgroup analyses and capture diverse experiences. Parents were contacted by the NIHR Bioresource via a personalised email and letter, followed by a phone call within two weeks. Invitations were sent in batches, prioritising older children. Each participant received a £15 gift card upon study completion.

### Study Design

This study used a convergent parallel mixed methods design to generate a multi-faceted understanding of parental perspectives, and to allow comparisons with clinical and non-clinical cohorts (30). Data integration occurred at multiple phases, with qualitative data given primacy alongside demographic data for group analyses. Quantitative measures were collected one day to a week before interviews; qualitative analyses were completed first, followed by statistical analyses. This method was chosen to inform data interpretation and triangulation.

### Measures

Parents completed questionnaires and provided demographic information online via Qualtrics™. Supplementary Table 1 lists questionnaire measures, internal validity metrics and scoring information. Questionnaire measures were selected after consultation with clinicians and researchers, and with a hospital parents’ group. Individual in-depth interviews (average length 95 minutes, range 46 min. – 2h 46 min.) were conducted via Zoom Video Communications (https://zoom.us/) with 91 parents by the primary researcher (H.D.) and recorded for transcription and analysis. A semi-structured interview guide, based on Aldridge et al. (31) was applied to elicit information about psychosocial impact of trio rGS, from which the questions on support are described here (Supplementary Table 2). Interview questions were piloted with parents of children with complex health conditions from two families (one mother, one father).

### Questionnaire analyses

Analyses were conducted using IBM SPSS Statistics (version 28.0). Where permitted by the scoring manual, we used mean imputation to estimate missing values; otherwise pairwise deletion was applied, indicated by the number of individuals in reported analyses. Distributions were explored for each variable, and parametric tests were appropriate.

Peregrin* group scores on outcome measures were compared with UK population norms (32,33). Where UK cohort data was unavailable, other non-clinical samples were utilised (34,35). Effect sizes were calculated as the difference between Peregrin* mean and comparison sample mean, divided by pooled standard deviation (36). Diagnosis and No Diagnosis sub-groups were compared using independent samples t-tests. To compare mothers’ and fathers’ well-being, within-couple analyses were carried out. Pearson correlations were used to compute bivariate associations and paired-samples t-tests or chi-squared (McNemar’s) were used to compare within-couple differences in means. Bonferroni corrections were applied for multiple testing.

### Interview Analyses

Interviews were transcribed verbatim, and all transcripts were checked for accuracy, with 68% reviewed by two researchers. Analyses were conducted in NVivo (Version 12) and in Microsoft Excel, using inductive and deductive content analysis (CA) (37,38), and framework analysis (FA) (39). Participant responses were coded line-by-line across all transcripts. The codebook for support sources, and framework for support group use, were developed from existing literature and from themes derived from close reading of 20 transcripts, using inductive and deductive approaches, and then applied to the entire data set. Transcripts were independently double coded by research assistants (S.R., M.B.), using the codebook and framework, and discrepancies resolved by consensus. All quotes have been anonymised and lightly edited for clarity, with an ellipsis in square brackets […] indicating that text has been shortened.

Support levels were categorised with four response options: “no support,” “minimal,” “sufficient,” or “high level.” Responses were dichotomised (none/minimal cf. sufficient/high) for comparison between Diagnosis/No Diagnosis (Dx / NDx) and Fathers/Mothers (F / M).

Broader responses were analysed thematically. Parental engagement with support groups was categorised based on overall use, current use, group type (e.g., condition-specific social media group, peer messaging group, rare disease charity), and perceived benefits and drawbacks. Single episodes of support group contact, independent online information-seeking, and unspecified support sources were excluded (40).

## RESULTS

### Cohort characteristics

755 parents were contacted (22 returned to sender). Of 192 respondents (25.5%), 139 expressed interest, 52 declined, 1 was ineligible, and 96 consented to participate. Reasons given for non-participation were time constraints and family commitments. Participants (32 couples; 32 parents participating alone) were parents of 67 affected children, representing 63 families (including 6 parents of 4 deceased children, and 4 sibling pairs). 91 parents participated in an interview; 5 parents completed questionnaires only. Parental self-reported demographic and health characteristics are described in Supplementary Table 3. Fathers’ (*n* = 37) mean age at interview was 41.0 years (*SD* = 5.6, range 31 - 53); mothers’ (*n* = 56) mean age at interview was 38.7 years (*SD* = 5.4, range 29 - 55).

Time since rGS at interview ranged from 1 to 5 years (mean 3.5 years, *SD* = 1.0); 57% of families had received a genomic diagnosis by the time of participation in Peregrin*. Children (30 male, 33 female) were aged 2 - 15 years (mean 5.6 years, *SD* = 2.7, excluding 4 deceased children) at the time of interviews. All children lived at home, with mean 2.14 (range 1 - 5) children per household.

Children’s clinical conditions and genetic diagnoses were heterogeneous, and often rare or ultra-rare. The most frequently affected body systems were nervous system (n = 41), digestive and respiratory systems (n = 35 each), cardiovascular system (n = 34) and growth (n = 27), i.e. many children had multi-system disorders. Children’s Health-Related Quality of Life (HRQL, PedsQL™ V4.0 Generic Core Scales) was significantly impaired relative to population norms for overall impact, physical functioning and psychosocial functioning (Supplementary Table 4).

### Questionnaire results – Family well-being and parents’ mental health Comparison to general population norms

Table 1 compares Peregrin* parents’ well-being indices with UK population norms (or equivalent non-clinical samples). Significant differences were observed in PedsQL™ Family Impact Module (PedsQL™ 2.0 FIM) total scale and several parent HRQL subscales (36). No differences emerged for parents’ physical functioning, family functioning total score, or family relationship scores. Life satisfaction scores remained within the average to high range and did not differ from norms (35,41). Table 2 reports Hospital Anxiety and Depression Scale (HADS) (42) comparisons - mothers reported significantly higher levels of anxiety (*d* = - 0.34) and depression (*d* = -0.28) compared to UK norms, while fathers did not.

**Table 1.** Family Well-Being: Comparison of Peregrin* Cohort to Population Norms.

| Scale | Population Norms *M*<br>(*SD*)<br>N variable | Total Cohort *M*<br>(*SD*)<br>N variable | Difference | Effect<br>Size |
| --- | --- | --- | --- | --- |
| **Family Impact<sup>1</sup>** | Non-clinical *N in*<br>*italics*<sup>a</sup> | **FIM** N = 86 | FIM | FIM |
| Total Impact | 70.8 (14.5) 726 | 61.7 (20.8) | 9.1 | 0.60 *** |
| **Parent HRQL** | 69.4 (15.5) 809 | 64.5 (20.7) | 4.9 | 0.30 * |
| Physical | 64.9 (17.4) 901 | 64.7 (22.3) | 0.2 | 0.01 |
| Emotional | 67.6 (17.9) 897 | 62.2 (23.9) | 5.4 | 0.29 * |
| Social | 74.4 (19.1) 896 | 62.7 (26.2) | 11.7 | 0.59 *** |
| Cognitive | 73.5 (18.6) 871 | 67.9 (22.1) | 5.6 | 0.30 * |
| Communication | 81.9 (17.7) 894 | 57.4 (26.9) | 24.5 | 1.31 *** |
| Worry | 78.1 (20.1) 876 | 50.6 (27.9) | 27.5 | 1.32 *** |
| **Family Functioning** | 65.5 (18.5) 870 | 63.6 (25.3) | 1.9 | 0.10 |
| Daily Activities | 63.2 (22.5) 901 | 51.0 (27.9) | 12.2 | 0.53 *** |
| Family Relationships | 67.0 (19.4) 881 | 71.2 (23.2) | -4.2 | -0.21 |
| **Subjective Well-Being<sup>2</sup>** | 23.46 (6.86)<sup>4</sup> | SWLS N = 94<br>24.5 (6.4) | SWLS<br>-1.04 | SWLS<br>-0.15 |

\*\*\* $p < 0.0001$ ; \*\* $p < .001$ ; \* $p < .05$ . Effect sizes are designated as small ( $d \sim 0.2$ ), medium ( $d \sim 0.5$ ), or large ( $d \sim 0.8$ or more).
*Note.* Higher values equal better health-related quality of life, family functioning, and higher subjective wellbeing. FIM = Family Impact Module; HRQL = health-related quality of life; SWLS = Satisfaction with Life Scale. <sup>1</sup>Data from Medrano et al., 2013. <sup>2</sup>Data from Siedlecki et al., 2008.
<sup>a</sup> Ages 18 - 39 ( $N = 200$ ) are reported to match the mean ages in the Peregrin\* sample of 32.9 for mothers (range 29 - 55) and 35.7 for fathers (31 - 53); the mean and standard deviation for ages 40 - 59 are 22.37 (7.29) ( $N = 310$ ).

**Table 2.**
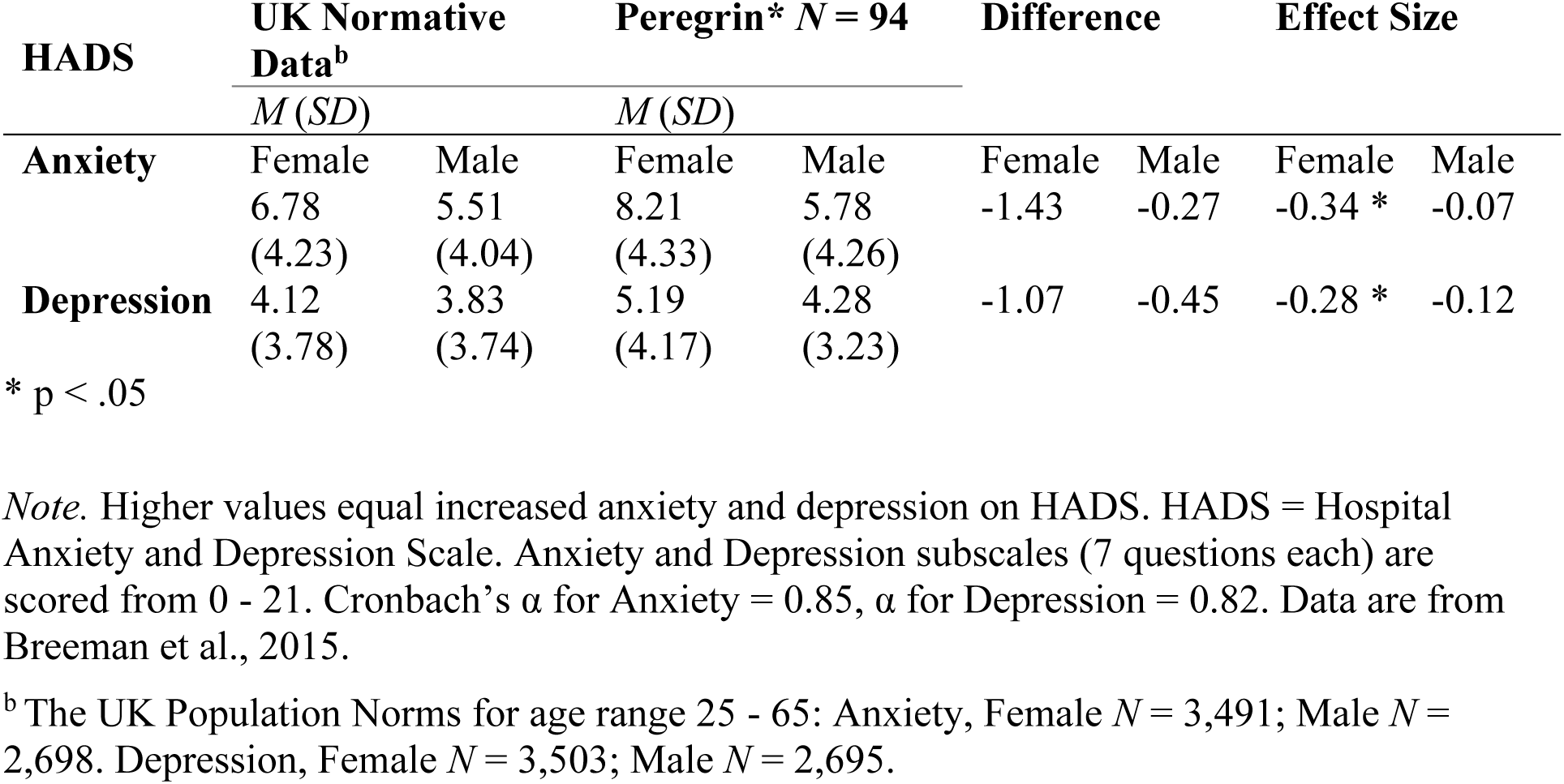
Parents’ Mental Health: Comparison of Peregrin* Cohort to Population Norms.

### Associations with diagnostic group

Parents in the Diagnosis group reported significantly higher total family impact, and expressed more worry, compared with the No Diagnosis group (Supplementary Tables 5 and 6). These two groups also differed significantly on Total Family Function, impacting on Daily Activities to a greater extent than Family Relationships. There were no group differences in anxiety, depression, or life satisfaction.

Children in the Diagnosis group had significantly lower HRQL compared to the No Diagnosis group on the Paediatric Quality of Life Inventory (PedsQL™ V4.0) (Total *t*(*84*) = 5.12, *p* < .001, one-sided), prompting us to re-examine well-being differences. *Post hoc* one-way analysis of covariance showed no significant difference in FIM total scores between Diagnostic groups (*F*₁, ₈₃ = 3.74, *p* = .057) but a significant covariate effect for child’s HRQL (*F*₁, ₈₃ = 80.72, *p* <.001). This indicates that parents’ psychosocial functioning may be explained by the complexity of children’s conditions, rather than the presence or receipt of a genomic diagnosis.

### Comparisons between fathers and mothers and associations within couples

Within-couple comparisons (Supplementary Table 7) confirmed that mothers reported significantly higher anxiety symptoms than fathers, while parents’ depression levels did not differ as measured by HADS. Mothers did, however, report more emotional difficulties on the FIM Emotional Functioning subscale. Mothers also reported greater worry regarding their child’s medical treatments, side effects, others’ reaction to child’s condition, its impact on other family members, and on child’s future. No significant differences between mothers and fathers emerged in Social Functioning or Communication about their child’s health and family situation.

Bivariate analysis of maternal and paternal variables showed a moderate within-couple association for Worry (Supplementary Table 7), but not for parents’ HRQL scores, mental health symptoms or total family impact scores. Instead, each parent’s FIM scores correlated strongly with their own HRQL (*r₍₃₀₎* = .949, .960, *p* < .001 for mothers and fathers, respectively), highlighting the individual nature of physical and mental health, and appraisal of family impact.

### Interview findings – understanding parents’ support

#### Sources and Perceptions of Support

Participants defined support in their own terms and named varied sources of primary support. Figure 1 presents the five most prevalent sources by parent gender (A) and by child’s diagnostic status (B), inspired by Gise et al. (43). In total, 19 categories of support were identified (Supplementary Figure 1). Table 3 summarises and illustrates the key qualitative findings on parental perceptions of support. Parents with adequate support cited emotional, informational, instrumental, esteem, and companionate support from both formal sources (e.g., hospital staff, counsellors, GPs, schools, hospices) and informal networks (e.g., spouse, parents, friends, colleagues). Some parents relied on a few key sources or were self-sufficient, knowing where to seek help when needed:

**Figure 1.**
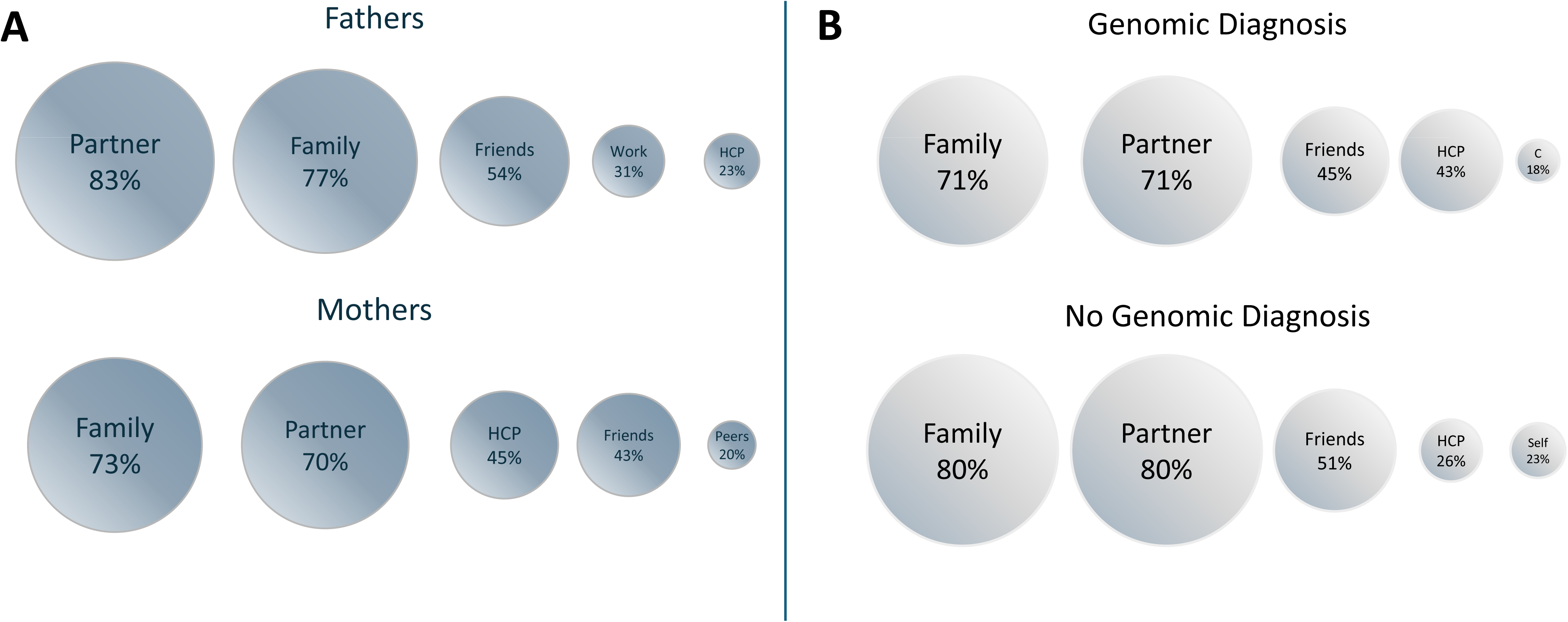
Five Most Prevalent Sources of Support by **(A)** Parent Gender, and by **(B)** Child’s Diagnostic Status *Note:* Abbreviations HCP = healthcare professionals, C = Counselling / Mental Health Support, Self = Self-Reliant.

**Table 3.** Key Qualitative Findings on Parental Perceptions of Support.

| Main findings | Illustrative quotes |
| --- | --- |
| **Parents evaluated support adequacy through the lens of personal, child, and family needs** | *Well, we're all functioning and surviving and fine; so, I guess we have sufficient support. INT33, M, Dx*<br>*I suppose when we think about support where our kids are concerned, we think about them being able – well, for [affected daughter] - her being able to access the things that she needs in order to have a good life and how well we can provision that. And we rely very heavily on support. INT4, M, Dx* |
| **Parents sometimes lacked emotional support due to differing coping styles or fear of burdening others** | *You can talk to your family and your mum as much as you like, but they don't know, they're not experts. [...] And yeah, it's like dealing with grief, isn't it? Yeah, everybody deals with it differently. And I yeah, I tried to say things to him [husband], but it was too much; it's too overwhelming for him to hear the worst end of the spectrum or what could potentially be the future. INT25, M, Dx* |
| **Peer parents became an important source of support for many mothers** | *[I turn] to another Mum who's got a child with the same diagnosis, for the most part, yeah, and she's somebody I've met through social groups, and we've met up several times, actually. But she will be my first point - for the most part. If it was something obviously very medical and urgent, then my son's liver team and the clinical nurse specialist there and yeah. So, I guess her and then the clinical nurse specialists are my saving graces every day. INT84, M, Dx* |
| **Parents' and child's support needs and support provision changed over time** | *Uh, sufficient now, definitely. Like now I've got the 12 hours and now he's at school - the special school - yeah, I'll get a break. INT32, M, Dx*<br>*Yeah, I'd say it's sufficient support, only because at the moment it's all in medical terms. INT27, M, Dx*<br>*Well, at the minute there's no support, no. Well, in a sense we don't have any support from the system as such, or from managers, or from the doctors. There's no support in terms of the mental [health] support. INT15, F, Dx*<br>*I suppose we're just perhaps [at] a bit of a crossroads of what the... how the support that we've got around us... is it going to be adequate enough to get [affected son] to the next phase of his life? which is sort of going into* |
|  | *secondary school and things like that. Yeah, I think there's always room to add more support. INT25, M, Dx* |
| **Parents utilised a variety of sources of support** | *We know that we can contact the hospital if we need anything, and we'll get a response. We've got school that we can call on. We've got parents. INT8, M, Dx* |
| **Parents' willingness, need, or capacity to seek or accept support differed – but parents expected support to be offered to all individuals regardless** | *I've got a high level of support. It's whether I ask for it is the big question. INT87, M, Dx*<br>*I find it very difficult having to be reliant on so many people. I really, really dislike it, like <u>really</u> dislike. I hate being dependent on other people, and I've never had to be, and I really hate it. I mean, I'm at the mercy of so many people! INT62, M, NDx*<br>*I feel that I personally I'm a person I like to find out as much information as I can. So, I do a lot of reading [...] At times it feels isolating and it's overwhelming [...] And so, I think I've sought some solace in being able to reach out and get a hold of doctor [specialist in son's rare condition] and his team, they're very responsive via e-mail. [...] and I feel going forward that that's going to be a really useful avenue to keep on with because I feel like those guys are [in] a better place to answer some of these difficult questions. INT28, M, Dx*<br>*What could make it better is to guide us as parents on the various steps, things that you should do: like the DLA, the EHCP, which is a jungle in itself. And in the beginning, even simple things like: how do you request prescription medication? We had no idea about anything. INT52, M, Dx* |
| **Parenting a child with CMC was experienced as a lonely journey without “right” support** | *I think it was just about connecting with other families who have gone through similar things. I think at the time I was feeling a bit alone, like all my friends' children were healthy and yet hadn't... didn't know anyone who had a similar experience. INT83, M, NDx* |
| **Getting or finding support was a “battle” for several families** | *It is down to us to do - to fight the fight, if you like. [...] in terms of physiotherapy and medical research, there is minimal support. INT7, M, Dx* |

*Note.* Abbreviations: CMC = child(ren) with medical complexity; INT followed by a number = interviewee number; M = mother, F = father; Dx = genetic diagnosis, VUS = variant of uncertain significance, NDx = no genetic diagnosis identified.

> *“Whenever I’ve had any concerns, and I’ve gone to the consultant, they’ve come straight back to us.” INT13, F Dx*

> *“We have a very close strong family base who we can rely on to keep us out of any trouble.” INT22, F, NDx*

> *“If I have concerns, I feel that I have people that I can talk to. […] I’ve sought some solace in being able to reach out and get a hold of doctor [expert in child’s condition] and his team.” INT28, M, Dx*

Partners and family were similarly important to most parents, while healthcare professionals were a more frequent support source for mothers than for fathers. Many parents reported a lack of emotional support from their extended family. In particular, grandparents were often perceived as either unable to understand their situation, emotionally distant, or in need of protection from distress. Some parents also lacked emotional support from their spouse/partner due to differing coping styles or fear of burdening an already overwhelmed partner:

> *“It’s mostly me and my husband that talk about things and I’ll speak to my sisters, and I speak to my mum […]. But […] sometimes I think they don’t really understand how she is.” INT2, M, Dx*

> *“I’m the man of the house sort of thing and, you know, you gotta let it wash over you sort of, and I don’t wanna bleat. […] There are times I’m thinking - I can’t talk to her anymore about it, because she does so much all the time!” INT5, F, Dx*

For fathers, friends and the workplace were key sources of emotional and social support, providing opportunities for venting, distraction, and self-mastery. Some employers offered flexible working arrangements, which helped to reduce stress related to childcare, medical appointments and financial pressures, while a few provided counselling services. In contrast, only 5.4% of mothers cited work as a source of support, possibly reflecting lower workforce participation (69.5% vs. 100%) and higher part-time employment rates (65.9% vs. 5.5%).

> *“Through the whole thing, we continue to both work, and we had very flexible and sympathetic employer.” INT6, M, Dx*

Mothers valued friends and peer parents for emotional, informational, and practical support, though many lacked time or energy for socialising, and some lost friendships due to differing parenting experiences. For some, peers (parents of children with the same condition or other health conditions/disabilities) became new closest friends:

> *“So, we’ve been - me and the mum - become friendly and we’re helping each other out, because there’s things that she knew, and I didn’t know.” INT65, M, NDx*

### Satisfaction with Support

74.7% parents reported “sufficient” or “high” levels of support, while 25.3% reported “no” or “minimal” support, with 6 parents giving mixed responses, which were averaged for the analyses (illustrative quotes, Table 4). Parents of children with a genomic diagnosis were much more likely to report insufficient support (36%) compared to those without a genomic diagnosis (6%). The differences between fathers’ (31%) and mothers’ (21%) satisfaction with support were small.

**Table 4.** Key Qualitative Findings on Parents’ Satisfaction with Support.

| Main Findings | Illustrative Quotes |
| --- | --- |
| **Satisfaction with level of support:** |  |
| **“None” or “Minimal”** | *At the time [of rGS] I would have said “sufficient”; now I would say “minimal”, because it's very much: “deal with it on your own now”. [...] it's very hard for them to keep track of everyone and to do everything and there's a lot more problems nowadays with different diagnoses and stuff that weren't available back in the day. But yeah, I do still feel there's very much a case of, yeah, the NHS is very stretched and they're not able to offer the services that they perhaps would like? INT70, F, VUS* |
| **“Sufficient” to “High”** | *Again, specialist wise [affected daughter] is pretty lucky. She's in good hands and she's got the right people in place, and then, like I say, support-wise for me and [husband], you know, it's not necessarily the quantity we have, but it's the quality that we have, and we know that there is always someone there. [...] I think we sometimes feel like we're imposing or, you know, it should be something we should be dealing with ourselves anyway, but you know, we get told off quite a lot [...] for obviously not asking for enough help. But you know, that's just how I think me and [husband] are. INT52, M, Dx* |
| **“Mixed”** | *Medically I have high support. Personally, I don't have any support, no. Medically, I have, I've got high support in terms of her die[t], like you know, the sleep studies that she gets, the OT, the physiotherapy - all that is pretty high - the nutritionist; and they're very responsive. So, I really appreciate that, but personally, I haven't got any support in terms of the social net or it's like, you know, the backup. I haven't got that. INT12, M, Dx* |
| **Satisfaction with types of support:** |  |
| **Lack of instrumental support** | *Support in a way we don't have any support for him. Me and my husband. We both are managing him because he is dependent for all the basic things. [...] We have emotional support from our families, but all are in [another country]. INT16, M, Dx* |
| **Lack of emotional support and access to mental health resources** | *I mean the GP is fairly useless, so it's just a case of relying on your support network. I rely on my work colleagues quite a bit to vent and I can always turn to my mum if* |
|  | *anything really gets to me, 'cause I don't like to burden the wife with it, she's got her own issues. INT51, F, Dx*<br>*So, I think with a diagnosis like that, counselling would have been incredibly useful. Part of me thinks that having some supports in how to deal with the diagnosis, someone that knows the condition quite well, someone to say: "There will still be good days," and, "this is the support that you will have along the way," [...] Because when you receive a diagnosis like that, you don't look at the near future, you look at the whole life. INT14, M, Dx* |
| **Lack of formal support and service provision, at time of diagnosis and across illness trajectory (including death)** | *Ultimately, you think when you got - when I got the test results even - okay, we're gonna get people help us now and, you know, they're gonna help us on our journey and do this, do that, and do the other, but we always seem to be fighting every single minute and every single appointment for [affected child]. INT5, F, Dx*<br>*I never was given information about the support groups, so I wouldn't even know where to start looking. INT77, M, Dx*<br>*So, I actually found her support myself by going to her school and getting it. But I do think that there needs to be something done with siblings because actually there is a - that is a gap. It's really hard just for young children to - they don't understand. INT90, M, Dx, Deceased* |
| **Unmet support needs, coupled with reluctance to burden others, and a preference for self-reliance** | *I mean there is an old saying that you can find it easier to talk to strangers. I found it easier to talk to you right now about my feelings than I would do my own Mum and Dad or my sister or anyone that I'm close to. Because the people you're close to, you know, they've got their own issues and they've got their own problems sort of thing you don't want it, like I said with [wife], you don't want to burden them with what's going on. INT5, F, Dx*<br>*I'm very conscious of the fact that my mum suffers with the same [mental health] issues that I suffer with, and so I tend to pick and choose if I talk to my mum about things. INT87, M, Dx* |

Parents reported receiving “no” or “minimal” support when either multiple support needs (e.g., medical, practical, and emotional) were unmet, their needs were acute (e.g., mental health deterioration, change in child’s health), available provision failed to meet expectations, the support was granted as the result of parental “*battling*” or “*fighting*”, or parent felt emotionally and socially isolated:

> *“No one’s spoken to us about it, no one has given us any information on it, and the new doctor - the Geneticist, he didn’t seem to know anything.” INT30, F, Dx*

> *“Actually, I recently went to a private therapist because I thought I had hit my ceiling. I thought I was at breaking point.” INT14, M, Dx*

> *“You could go and talk to any sort of counsellor or whatever, but unless they’ve actually experienced what it is like to live with a child with these really challenging points of view, it doesn’t matter how many - you know - how well qualified this counsellor is. If you just don’t get it, there’s no point.” INT20, F, NDx*

> *“With the genetic? Hell, “no” - not really. It’s just having to kind of fight for the speech and language and the physio. I kind of feel like I’m just getting fobbed off all the time.” INT60, M, VUS*

> *“We’re kind of isolated really, you know. And I think probably if I had, if I could do more things with friends, then maybe I wouldn’t feel so… maybe isolated.” INT2, M, Dx*

Key elements of insufficient support included: (i) lack of practical home-based support, often due to distant or elderly family members, or lack of information about social care options; (ii) feeling emotionally unsupported by healthcare providers, and experiencing long wait times for mental health assessments or family counselling due to lack of psychological service provision; (iii) limited organisational support and service co-ordination, requiring significant effort to obtain necessary health and educational services for their child; (iv) unmet informational or emotional support needs, with uncertainty about where to seek help, reluctance to burden others, or a preference for self-reliance. Some reported adequate medical support for the child but unmet parental support needs, or changes in support over time. Despite individual and gender differences in parents’ willingness, need, or capacity to seek or accept support, parents expected support to be offered to *all* parents.

### Engagement with Support Groups

Parents accessed diverse support groups including online condition-specific, general (inter)national rare disease organisations, disability-related networks, and in-person specialist baby/toddler groups. Online platforms, particularly Facebook groups, were the most frequently used, offering flexibility and accessibility, although availability varied by condition, prompting some parents to create new groups, tailored to their child’s diagnosis. Mothers engaged in support groups more than twice as often as fathers (78.2% vs. 34.3%). However, both parents acknowledged the value of such groups, often supporting other individuals/families themselves. In most couples, at least one parent participated, allowing the other to benefit indirectly.

> *“My partner is member of a group - the [syndrome] group on Facebook. […] So, we… well, [wife] more so… uses that just to keep abreast of the whole, you know, the condition and, […] with research and all that sort of stuff really.” INT24, F, Dx*

Parents identified both benefits and challenges to support group involvement. Membership facilitated shared experiences and offered guidance on managing the child’s condition. Emotional and informational support helped to reduce parental isolation and fostered a sense of belonging, especially when emotional support from family and friends was lacking. Practical advice focused on therapies, diet, applying for disability benefits, schooling, equipment, disability access in social spaces, and travel.

> *“Like she’s nonverbal, so I never really know… Any epilepsy sites are helpful because for people who’ve got it, they can tell you how they feel like before they have one and after. […] A lot of times I’ll go on there to see what people think about various medications.” INT18, M, Dx*

> *“People have said before: “Oh, do you not want to speak to other parents?” I really haven’t wanted to speak to other parents because I really wanted us to manage our situation as it is for us as a family and for [daughter] as it is for her. I didn’t want to take on other people tales of woe and listen to how awful everything is. Anyway, having since joined this Facebook group, it’s been great in terms of knowledge and information sharing and knowing what else is going on.” INT7, M, Dx*

However, support groups were described as a “double-edged sword,” providing valuable resources but also heightening anxiety about the child’s future. They were unsuitable for parents who described themselves “unsociable”. Additionally, not all parents identified as “support group users”, preferring to use terminology such as “parent forums” or “networks” instead. Some parents disengaged due to emotional distress, lack of shared experiences, or changing needs over time.

> *“[…] I did find a page on Facebook of all places - a [syndrome] Facebook. And there are people all over the world who’ve got kids with the same diseases […] And people put videos of their kids on there who are the same age as [child], and […] it’ll just give you a bit of a boost. And then you see the other side of the flip story, where they’ve got a little lens on there who are a lot worse than [child], and it hits home sort of thing.” INT5, F, Dx*

> *“Now on this group - I remember signing up to it, and I couldn’t believe what I was seeing. It was quite scary. And yeah, I’m still on the group, but they don’t like me on there.” INT10, M, Dx*

> *“Who’s got the time to go and attend support group? My day is full on. I started 7:00 o’clock and I ended 11:00 o’clock.” INT12, M, Dx*

Parents’ engagement with groups fluctuated / evolved: some parents withdrew after initial participation (13/55, 23.6%), while others joined after an adjustment period of months to years, as their needs changed. Reasons for disengagement included returning to work and time constraints, variability within condition-specific groups causing distress, and lack of shared experiences. Some parents were excluded due to eligibility criteria (e.g., bereavement groups for foetal vs. infant loss). Others found peer support relationships unfulfilling, outgrew the need for support, or encountered group discontinuation.

> *“There’s Facebook groups to join: dads’ groups, bereavement groups […] - and I have joined a couple of them. [Wife] joined one, and she said it just feels like it’s just little women bitching. […] I don’t go on it anymore.” INT91, F, Dx*

> *“I tried to join a group actually through SAND’s and I wasn’t allowed.” INT90, M, Dx*

After hospital discharge, many parents found it difficult to access peer support and missed out on early socialising and intervention opportunities. Barriers included lack of information or absence of suitable groups, time constraints, work commitments, or a preference for alternative support sources such as health professionals, family or friends. Noticeable differences in one’s child’s development within non-specialist (mainstream) baby/toddler groups, and lack of father-specific support groups further hindered participation (Supplementary Table 8).

## DISCUSSION

The Peregrin* study identified vulnerability and variability in well-being and quality of life among parents of children who had received early rGS - the well-being of the cohort was compromised, and support needs were not fully met. Parents faced barriers to support, reinforcing the need for timely, inclusive, accessible and varied resources for parents of children with complex medical conditions (44). Contrasts in needs emerged between diagnostic groups, with parents of children with rare genetic, medically-complex conditions especially at risk of emotional distress. A third of these parents reported minimal or no support, contrasting with the expectation that obtaining a genetic diagnosis will enhance access to support and improve well-being (24,45). This is consistent with previous findings that parents in receipt of (rapid) genomic sequencing results, and those caring for children with (rare) complex conditions, frequently experience caregiver stress due to emotional, social and financial pressures, but often lack the right support at the time of highest need (6,7,46). Across follow up studies, a subset of parents reports ongoing high anxiety and/or depression (14,47,48). This mismatch between the needs of most-impacted families and low support availability necessitates a holistic and dynamic approach, ensuring sustained family support that addresses both child-focused medical care and parental well-being.

An important element of this study was the comparison between mothers’ and fathers’ needs and support experiences. Consistent with previous findings (49), mothers reported above general population levels of anxiety, worry and emotional difficulties, whereas this was not seen at group level for fathers. Within-couple analysis indicated independent mental health symptom scores between parents, emphasising the need for individualised psychological interventions for parents. Fathers, similar to those of typically developing children, described stress rather than mental health difficulties and often questioned their entitlement for professional support, prioritising the needs of their partner (50). The differences between parents’ adjustment and adaptation deserve careful consideration in future research (20).

Parents defined support based on their specific needs and assessed its adequacy in relation to their child, themselves, and their families. Both formal and informal support networks were described as crucial in fostering a sense of belonging and facilitating shared experiences among families. As observed previously in families managing medical complexity and genetic conditions (43,51,52) perceived (functional) and actual (structural) parental support changed over time. Informational and emotional support was most critical during the diagnostic phase and at key transitions, while instrumental (e.g., respite care) and social support became essential for long-term family well-being. Despite observable gender differences in support utilisation, there was considerable overlap, with some parents engaging in social networks typically associated with the opposite gender.

Family and partners were primary support sources for most parents, indicating the need to foster strong family relationships, particularly when under strain from a child’s diagnosis. However, partnership status alone is an unreliable proxy for social support (53). Parents who are heavily dependent on partners may require professional intervention when relationship strain or mental health concerns arise. While many parents reported satisfactory social functioning, a considerable minority experienced isolation and difficulties securing support. Parents with high self-reliance often viewed seeking support as a weakness, despite experiencing isolation and insufficient emotional connection. Primary caregivers would benefit from tailored interventions to maintain friendships and mitigate social isolation, especially during diagnosis and transitional periods (46,54).

It has been reported that rare disease diagnosis can facilitate access to condition-specific support groups (6), yet many parents in Peregrin* did not access support groups, emphasising the need for alternative sources of support. Some families preferred guidance from trusted healthcare providers rather than charitable organisations or peer networks. Additionally, not all social support was perceived as beneficial. Support groups could be distressing for parents confronted with potential deterioration or loss, were considered unsuitable by parents who described themselves as “*unsociable*” or “*self-reliant”*, or who preferred their child’s condition not to dominate their spare time and life experience. Thus, while support groups benefit many, they should be supplementary rather than the primary mode of support. Bolstering partner and family relationships should be a priority in support provision, while professionals, employers and friends also contribute to meeting families’ long-term needs (52).

### Limitations

The complexity and heterogeneity of conditions diagnosed within our study group may limit generalisability to less severe or more common genetic conditions. The study’s cross-sectional design precluded assessment of temporal changes, with potential biases of retrospective reporting, in the context of evolving adjustment. Despite a diverse cohort and data from both mothers and fathers, sample size limits quantitative analyses. There are also limitations of demographic representation - a high proportion of parents had degree-level education and were either married or cohabiting, and participants were recruited from a tertiary university hospital, potentially differing in access to healthcare and social services. While we followed the methodology adopted in non-clinical samples, maximising participation and comparisons by including both mothers and fathers, non-independence within the dataset introduces a potential bias. Longitudinal research is needed to examine changes in parental support needs following a child’s diagnosis of a rare genomic condition. Studies should also investigate resilience and burnout in parents who exhibit high independence but face caregiving stress (55).

### Conclusion and Practice Implications

Based on Peregrin*, healthcare professionals can improve their support of parental well-being in several ways. First, practitioners can recognise the differential impacts of a child’s diagnosis on individual parents and validate each parent’s unique experience and role within their family. Second, self-reliant and highly independent parents may benefit from normalising support-seeking behaviours and sharing of research evidence linking support with improved well-being. Alternative terminology to “support” (e.g., networks, resources, or forums) may increase engagement (of fathers in particular), without affecting parental self-esteem. Third, support assessments and practical referrals should be integrated and their importance reinforced, benefiting families most in need of and least able to access support. Lastly, psychological support should be a core component of initial diagnostic and long-term care for families who are managing complex medical and genetic conditions. Our findings highlight the diversity of parental support preferences and the importance of offering flexible, timely, and tailored support options at the time of genomic testing and across the child’s illness trajectory, to accommodate families’ evolving needs and circumstances.

## Data Availability

Additional data supporting the findings of this study are available in the Apollo repository [https://doi.org/10.17863/CAM.121146], as part of the author’s PhD thesis. Deidentified data are available upon request and should be addressed to the corresponding author.

## Acknowledgements

We thank NIHR BioResource volunteers for their participation, and gratefully acknowledge NIHR BioResource centres, NHS Trusts and staff for their contribution. We thank the National Institute for Health Research, NHS Blood and Transplant, and Health Data Research UK as part of the Digital Innovation Hub Programme. The views expressed are those of the author(s) and not necessarily those of the NHS, the NIHR or the Department of Health and Social Care.

We thank Madeline Connell (M.C.), Charlotte Burdge (C.B.), and Kate Poulter (K.P.) for data cleaning and research support.

## Author Contributions

Conceptualisation: H.D., K.B., C.H.; Data Curation and Project Administration: H.D.; Funding Acquisition: H.D., L.R., D.H.R., C.H., K.B.; Formal Analysis: H.D., S.R., M.B.; Resources: H.D., S.R., M.B., L.R.; Supervision: C.H., K.B., S.O.C., S.A.; Writing-original draft: H.D., K.B., C.H., S.O.C.; Writing-review and editing: H.D., K.B., C.H., L.R., S.O.C., D.H.R., S.A., S.R., M.B.

## Ethics Declaration

This Peregrin* follow-up study with the parents from the Next Generation Children project (NGC, REC reference 13/EE/0325) was approved by the local research ethics committee (REC reference: 21/EE/0154, HVS/2020/3264, IRAS: 300380), by the NIHR Bioresource (NBR133), and by the Cambridge University Hospitals NHS Foundation Trust institutional review board (A095947). Written electronic informed consent was obtained from all participants prior to study commencement and archived. All data was de-identified. The study adhered to the principles set out in the Declaration of Helsinki.

