## Supplementary figures and images for "Fathers’ and Mothers’ Support Needs and Support Experiences after Rapid Genome Sequencing"

### Supplemental Figure 1

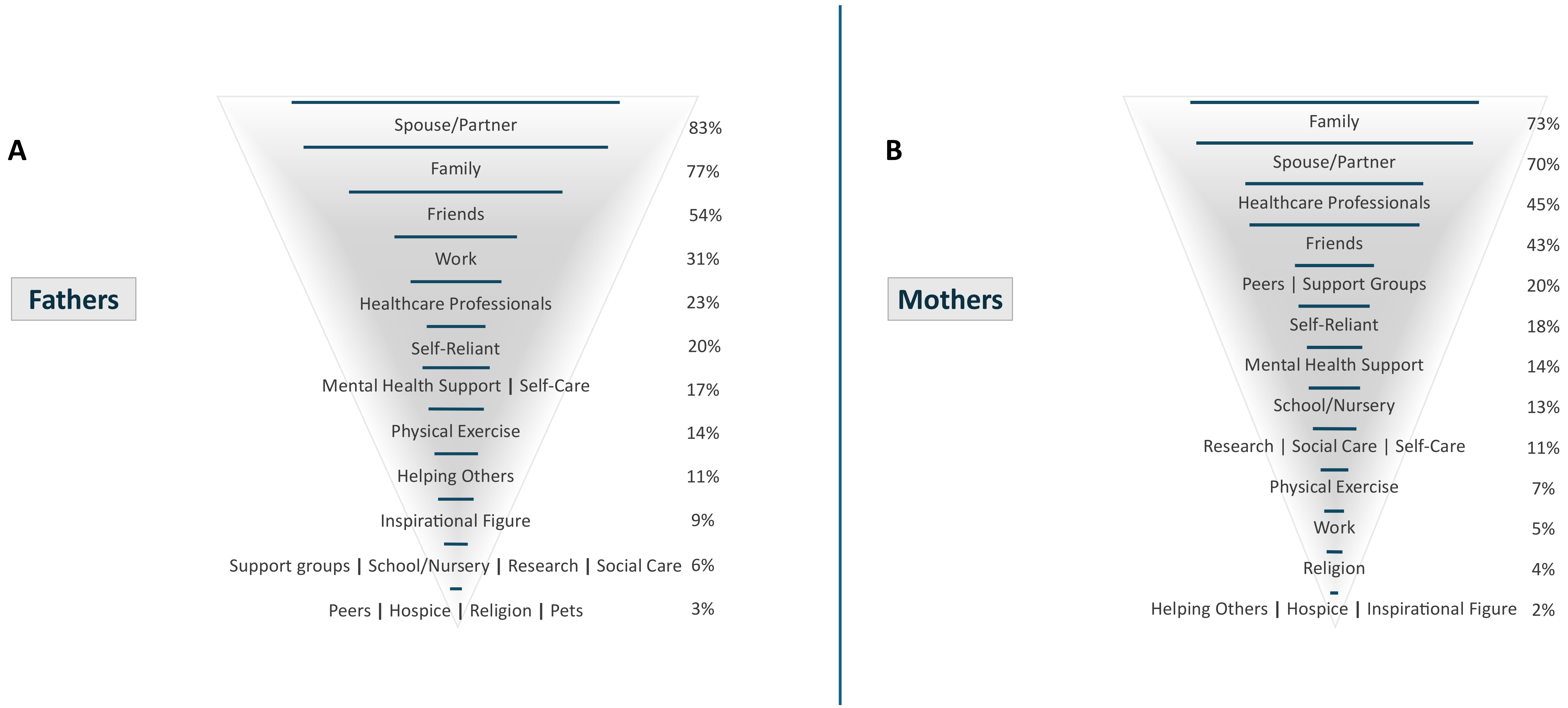
