## Supplemental Tables for "Fathers’ and Mothers’ Support Needs and Support Experiences after Rapid Genome Sequencing"

**Supplementary Table 1.** Questionnaire Measures

| Domain | Measure | Cronbach's alpha | Scoring notes | Reference |
|---|---|---|---|---|
| Parental quality of life and family functioning (FIM)* | 36-item PedsQL™ Family Impact Module (PedsQL™ 2.0 FIM) | Subscales α = 0.79 to 0.94<br>Total scale α = 0.97 | Parent QL is computed by averaging 20 items comprising Physical, Emotional, Social and Cognitive Functioning subscales.<br>Family Functioning is computed by averaging the 8 items in the Daily Activities and Family Relationships subscales.<br>Social Life subscale [a]<br>Communication subscale [b] | (30) |
| Child health-related quality of life (HRQL)* | PedsQL™ V4.0 Generic Core Scales; physical and psychosocial health scores | Subscales α = 0.73 to 0.95 | Higher score indicates higher level of functioning (less negative impact). Minimal clinically important difference 4.5 | (31) (32) |
| Anxiety and depressive symptoms (HADS) | Hospital Anxiety and Depression Scale | Anxiety α = 0.85, Depression α = 0.82 | non-cases (<7)<br>mild (8-10)<br>moderate (11-14)<br>severe (15-21) | (33) (34) |
| Subjective well-being and life satisfaction (SWLS) | Satisfaction with Life Scale | α = 0.89 | five-item scale scores 5-35; scores 20-24 indicate average satisfaction | (35) (36) |

*Parents of deceased children (n = 6) did not complete these measures.

| | |
|---|---|
| [a] FIM Social Life subscale (Cronbach's α = 0.90) | I feel isolated from others<br>I have trouble getting support from others<br>It is hard to find time for social activities<br>I do not have enough energy for social activities |
| [b] FIM Communication subscale (Cronbach's α = 0.79) | I feel that others do not understand my family's situation<br>It is hard for me to talk about my child's health with others<br>It is hard for me to tell doctors and nurses how I feel |

**Supplementary Table 2.** Interview Questions on Sources and Perceptions of Support

| Interview Questions & Prompts |
|---|
| **1. What are your primary sources of support?** (i.e. "first port of call") *Follow-up:* Did your support change with the results of WGS (or illness)? *Prompt:* Can you tell me a little more about how you find * supportive? |
| 2. **Do you feel you have enough support overall? (No, minimal, sufficient, high)** *Prompt:* Can you tell me about how you chose your response or what your thought processes were? |
| 3. **How do you feel about support groups? Have you joined any?** *Prompt:* Can you tell me a little more about how you use these support groups? *Or:* Can you tell me a little more about your thoughts and feelings about support groups? |

**Supplementary Table 3.** Parent Characteristics

| Parent Characteristics | N = 96 (%) | Genomic Diagnosis | Clinical Diagnosis |
|---|---|---|---|
| Number of Couples | 32 (66.7) | 20 (69.0) | 12 (63.2) |
| Number Participating Alone | 32 (33.3) | 18 (31.0) | 14 (36.8) |
| **Gender** | | | |
| Female | 59 (61.5) | 37 (63.8) | 22 (57.9) |
| Male | 37 (38.5) | 21 (36.2) | 16 (42.1) |
| **Current Age[1]** | | | |
| 25-29 | 1 (1.0) | 1 (1.7) | 0 (0.0) |
| 30-34 | 15 (15.6) | 5 (8.6) | 10 (26.3) |
| 35-39 | 34 (35.4) | 22 (37.9) | 12 (31.6) |
| 40-44 | 27 (28.1) | 16 (27.6) | 11 (28.9) |
| 45-49 | 13 (13.5) | 11 (19.0) | 2 (5.3) |
| 50-54 | 5 (5.2) | 2 (3.4) | 3 (7.9) |
| 55-59 | 1 (1.0) | 1 (1.7) | 0 (0.0) |
| **Ethnicity** | | | |
| White English, Welsh, Scottish, Northern Irish or British | 75 (78.1) | 46 (79.3) | 29 (76.3) |
| White European | 11 (11.5) | 7 (12.1) | 4 (10.5) |
| White & Asian | 1 (1.0) | 0 | 1 (2.6) |
| White & Black African/Caribbean | 2 (2.1) | 1 (1.7) | 1 (2.6) |
| Indian | 4 (4.2) | 4 (6.9) | 0 |
| Any other mixed or multiple ethnic background | 2 (2.1) | 0 | 2 (5.3) |
| Any other ethnic group | 1 (1.0) | 0 | 1 (2.6) |
| **Marital status** | | | |
| Married, or in domestic partnership | 82 (85.4) | 51 (87.9) | 31 (81.6) |
| Single (never married or never registered a partnership) | 12 (12.5) | 5 (8.6) | 7 (18.4) |
| Divorced | 2 (2.1) | 2 (3.4) | 0 |
| **Education[2]** | | | |
| Less than a high school diploma | 10 (10.4) | 10 (17.2) | 0 |
| High school degree or equivalent | 10 (10.4) | 7 (12.1) | 3 (7.9) |
| NVQ or equivalent | 16 (16.7) | 13 (22.4) | 3 (7.9) |
| Some college, no degree | 5 (5.2) | 2 (3.4) | 3 (7.9) |
| Bachelor's degree | 27 (28.1) | 16 (27.6) | 11 (28.9) |
| Master's degree | 16 (16.7) | 6 (10.3) | 10 (26.3) |
| Doctorate or professional degree | 11 (11.5) | 4 (6.9) | 7 (18.4) |
| **Employment status** | | | |
| Employed full time | 43 (44.8) | 25 (43.1) | 18 (47.4) |
| Employed part time | 29 (30.2) | 17 (29.3) | 12 (31.6) |

| | | | |
|---|---|---|---|
| Self-employed | 6 (6.3) | 4 (6.9) | 2 (5.3) |
| Looking after home or family | 14 (14.6) | 10 (17.2) | 4 (10.5) |
| Unable to work | 2 (2.1) | 1 (1.7) | 1 (2.6) |
| Unemployed & currently looking for work | 2 (2.1) | 1 (1.7) | 1 (2.6) |
| **Physical Health[3]** | | | |
| Very good | 38 (39.6) | 21 (36.2) | 17 (44.7) |
| Good | 40 (41.7) | 25 (43.1) | 15 (39.5) |
| Fair | 16 (16.7) | 11 (19.0) | 5 (13.2) |
| Bad | 1 (1.0) | 1 (1.7) | 0 |
| **Mental Health[3]** | | | |
| No | 67 (69.8) | 45 (77.6) | 22 (57.9) |
| Yes, in the past | 11 (11.5) | 4 (6.9) | 7 (18.4) |
| Yes, at present | 9 (9.4) | 6 (10.3) | 3 (7.9) |
| Both past & present | 8 (8.3) | 3 (5.2) | 5 (13.2) |

*Note*. [1] Age brackets are based on age groups used in the population normative data for the Hospital Anxiety and Depression Scale (HADS). [2] One missing in Clinical Diagnosis group (2.6%). [3] Parents were asked about any mental health conditions affecting mood, thinking or behaviour and requiring treatment or time off work.

**Supplementary Table 4.** Child Health-Related Quality of Life

| Scale | Population Norms *M (SD)* | Total Cohort *(SD)* | Difference | Effect Size |
|---|---|---|---|---|
| **PedsQL™** | UK Healthy *N = 665*[1] | **PedsQL™** *N = 86* | PedsQL™ | PedsQL™ |
| Total Impact | 84.61 (11.19) | 58.3 (24.1) | 26.31 | 1.98 *** |
| Physical Functioning | 89.06 (12.27) | 53.0 (35.2) | 36.06 | 2.18 *** |
| Psychosocial Functioning | 82.21 (12.67) | 61.3 (21.2) | 20.91 | 1.50 *** |

*** p < 0.0001; ** p <.001; * p < .05. Effect sizes are designated as small (d ~ 0.2), medium (d ~ 0.5), or large (d ~0.8 or more).

*Note.* Higher values equal better health-related quality of life and functioning on PedsQL™. PedsQL™ Total Impact scores are computed by averaging all items on the scale. Physical Health is computed by averaging Physical Functioning scores; Psychosocial Health by averaging Emotional, Social, and Nursery/School Functioning subscales. PedsQL™ = Paediatric Quality of Life. [1] Data is from Upton et al., 2005.

**Supplementary Table 5.** Comparisons Between Diagnostic Groups

| Scale | Genomic Diagnosis M (SD) | Clinical Diagnosis M (SD) |
|---|---|---|
| **FIM PedsQL™** | FIM N = 51 Parents[1] | FIM N = 35 Parents[2] |
| Total Impact | 58.1 (20.5) * | 67.1 (20.2) * |
| **Parent HRQL** | 62.4 (20.3) | 67.5 (21.2) |
| Physical | 63.0 (21.8) | 67.2 (23.1) |
| Emotional | 60.9 (24.5) | 64.0 (23.3) |
| Social | 60.4 (26.8) | 66.1 (25.2) |
| Cognitive | 64.8 (21.6) | 72.3 (22.5) |
| **Communication** | 54.9 (27.1) | 61.0 (26.5) |
| **Worry** | 42.7 (26.9) ** | 62.3 (25.4) ** |
| **Family Functioning** | 58.2 (25.8) * | 71.5 (22.5) * |
| Daily Activities | 42.1 (33.4) * | 63.8 (33.1) * |
| Family Relationships | 67.8 (24.7) | 76.1 (20.3) |
| **Child HRQL PedsQL™** | PedsQL™ *n = 51* | PedsQL™ *n = 35* |
| Total Impact | 48.6 (20.7) ** | 72.4 (22.0) ** |
| Physical Functioning | 37.3 (30.6) ** | 75.9 (28.4) ** |
| Psychosocial Functioning | 55.1 (19.8) ** | 70.3 (20.1) ** |

*** $p < 0.0001$; ** $p <.001$; * $p < .05$.

*Note.* Higher values equal better health-related quality of life and family functioning on FIM and PedsQL™. [1] 6 parents of deceased children did not provide data; [2] Two parents were excluded due to missing data.

**Supplementary Table 6.** Comparisons of Wellbeing Between Diagnostic Groups

| HADS | **Genomic Diagnosis**<br>**HADS & SWLS *N = 59*** | | **Clinical Diagnosis**<br>**HADS & SWLS *N = 37*** | |
|---|---|---|---|---|
| **Anxiety** | **Female** N = 37 | **Male** N = 21 | **Female** N = 22 | **Male** N = 16 |
| | 7.83 (4.42) | 6.38 (4.60) | 8.82 (4.21) | 4.93 (3.73) |
| **Depression** | 5.22 (4.36) | 4.57 (3.47) | 5.14 (3.96) | 3.87 (2.92) |
| **SWLS** | 23.8 (6.8) | | 25.2 (6.0) | |

All comparisons non-significant.

*Note.* Higher values equal increased anxiety and depression on HADS. Female (Genomic Diagnosis, 2 excluded due to missing data). Higher values on SWLS equal greater life satisfaction.

**Supplementary Table 7.** Comparisons Between and Associations Within Couples

| Scale | Mother M (SD) | Father M (SD) | M - F Correlations |
|---|---|---|---|
| | n = 30 | n = 30 | n = 60 |
| **FIM PedsQL™** | | | |
| Total Impact | 58.7 (20.9) | 66.2 (21.3) | .301 |
| **Parent HRQL** | 61.4 (21.1) | 69.6 (20.3) | .008 |
| Physical | 61.8 (22.8) | 68.7 (23.2) | -.039 |
| Emotional | 57.7 (23.7) † | 69.5 (23.9) † | -.057 |
| Social | 60.0 (28.1) | 69.8 (23.4) | .207 |
| Cognitive | 65.8 (23.8) | 70.7 (22.8) | .029 |
| **Communication** | 53.6 (24.0) | 61.4 (30.0) | .289 |
| **Worry** | 43.7 (28.6) † | 57.2 (26.2) † | .388 † |
| **Family Functioning** | 63.3 (26.2) | 65.0 (27.1) | .596 †† |
| Daily Activities | 47.5 (34.8) | 56.7 (36.8) | .586 †† |
| Family Relationships | 72.8 (24.7) | 70.0 (23.8) | .596 †† |
| **Child PedsQL™** | | | |
| Total Impact | 57.1 (24.5) | 60.3 (23.5) | .676 †† |
| Physical Health | 50.5 (34.1) | 57.7 (35.5) | .747 †† |
| Psychosocial Health | 60.8 (21.4) | 61.8 (20.4) | .534 †† |
| **HADS** | N = 32 | N = 32 | N = 64 |
| Anxiety | 8.1 (4.4) † | 5.5 (4.2) † | -.062 |
| Depression | 4.5 (3.7) | 3.8 (3.2) | .016 |
| **SWLS Total** | 25.1 (6.5) | 25.3 (6.4) | .363 † |

†† significant between mothers and fathers at <.001; † significant at 0.05 level.

*Note.* M - F = mother and father.

**Supplementary Table 8.** Parents' Views and Engagement with Support Groups

| Views and Engagement | Illustrative Quote |
|---|---|
| **Benefits and costs** | *I found a Facebook group for parents of children with [rare syndrome] which was terrifying and enlightening - would be in equal measures. I eventually come out of it because I didn't relate to a lot of it, because a lot of the families in there their children have been more severely affected by [syndrome].* INT25, M, Dx |
| **Being excluded due to a group's narrow remit** | *Well, when she was born, I went to all the brain injury trusts and every single one rebuffed me and said they couldn't help me because we didn't fall under their remit. And that was really awful as an experience, 'cause every single one rejected me. So, then it just… like you don't then want to go and ask for help, because it's hard actually being rejected.* INT62, M, NDx |
| **Lack of groups for fathers** | *The Facebook page [...] there's 1000 people on there, let's say 995 of them are mums, you know? And they're always talking to each other and that sort of thing, and there doesn't seem to be a lot there for the dads. […] It'd be nice to talk to some of the dads actually on there, but there are no dads on there.* INT5, F, Dx |
| **Support groups did not suit all parents** | *On Facebook [rare disease] there's this group there, from all around the world. I didn't get anything from it whatsoever. All the children are so different, and I haven't yet to meet someone in the UK that has this condition. Am I really one of those people that meet up with other mums with the same condition? In all honesty, no.* INT11, M, Dx |
| **Lack of time and energy to engage with support groups** | *I could join, I'm OK to join support groups, but the thing is, our day is so hectic, because when [affected daughter] comes back from school we've got lots of things around [daughter] to do. And we want to give her bath, and that's a big routine, and massage. Now we gotta do her medications* |

| | |
|---|---|
| | *and household chores and all that.* INT15, F, Dx |
| **Temperamental preferences affected support group use** | *I've not particularly felt the need to, to be fair. Like I say, I mean, you know, I am a bit of a miserable ****** and I do like a moan, but I generally think that I'm able to - I don't feel like I struggle to handle... So yeah, I would probably say the closest thing I have to a support group is when I'm moaning to people about things for me and then I'm sure that has its own help; but no, I've never felt the need for a support group.* INT76, F, NDx |
| **Discomfort with the language of framing parent/disability groups as "support groups"** | *I think support groups are really, really good. Really important. I've not joined any… Well, no, I suppose I have. I guess I don't see them as support groups in that way. I suppose depends maybe exactly kind of how you'd categorise them. [lists a rare diseases charity, Facebook group, local rare disease dance group, and disability play group] So, I suppose yeah, initially I wasn't thinking of those as 'support groups' because they're not put out there as a, "Oh, you're struggling. Join a support group." But they are very important networks, and I suppose that's where I get support from, you know, knowing that there is contact with other people who may be experiencing similar situations with their children.* INT38, F, Dx |
